# Deep Learning of Electrocardiograms Enables Scalable Human Disease Profiling

**DOI:** 10.1101/2022.12.21.22283757

**Authors:** Rachael A. Venn, Xin Wang, Sam Freesun Friedman, Nate Diamant, Shaan Khurshid, Paolo Di Achille, Lu-Chen Weng, Seung Hoan Choi, Christopher Reeder, James P. Pirruccello, Pulkit Singh, Emily S. Lau, Anthony Philippakis, Christopher D. Anderson, Patrick T. Ellinor, Jennifer E. Ho, Puneet Batra, Steven A. Lubitz

## Abstract

The electrocardiogram (ECG) is an inexpensive and widely available diagnostic tool, and therefore has great potential to facilitate disease detection in large-scale populations. Both cardiac and noncardiac diseases may alter the appearance of the ECG, though the extent to which diseases across the human phenotypic landscape can be detected on the ECG remains unclear. We developed a deep learning variational autoencoder model that encodes and reconstructs ECG waveform data within a multidimensional latent space. We then systematically evaluated whether associations between ECG encodings and a broad range of disease phenotypes could be detected using the latent space model by deriving disease vectors and projecting individual ECG encodings onto the vectors. We developed models for both 12- and single-lead ECGs, akin to those used in wearable ECG technology. We leveraged phecodes to generate disease labels using International Classification of Disease (ICD) codes for about 1,600 phenotypes in three different datasets linked to electronic health record data. We tested associations between ECG encodings and disease phenotypes using a phenome-wide association study approach in each dataset, and meta-analyzed the results. We observed that the latent space ECG model identified associations for 645 (40%) diseases tested in the 12-lead model. Associations were enriched for diseases of the circulatory (n=140, 82% of category-specific diseases), respiratory (n=53, 62%), and endocrine/metabolic (n=73, 45%) systems, with additional associations evident across the human phenome; results were similar for the single-lead models. The top ECG latent space association was with hypertension in the 12-lead ECG model, and cardiomyopathy in the single-lead ECG model (p<2.2×10^-308^ for each). The ECG latent space model demonstrated a greater number of associations than ECG models using standard ECG intervals alone, and generally resulted in improvements in discrimination of diseases compared to models comprising only age, sex, and race. We further demonstrate how a latent space model can be used to generate disease-specific ECG waveforms and facilitate disease profiling for individual patients.

## INTRODUCTION

The modern electrocardiogram (ECG) utilizes waveform data generated from surface electrodes to represent cardiac activation and impulse conduction.^1^ Introduced in the early 1900s, the original ECG was primarily used for arrhythmia detection, but its diagnostic utility expanded rapidly to include identification of coronary artery disease and other cardiac structural abnormalities.^2,3^ It has now become clear that non-cardiac diseases, from electrolyte derangements to central nervous system pathology, also cause characteristic changes in the ECG waveform.^4-7^

Recent advances in machine learning have revealed that the ECG contains diagnostic and prognostic information that extends beyond traditional clinical interpretation.^8-11^ Low-dimensional representations of ECGs constructed from deep learning models can detect cardiac diseases such as left ventricular dysfunction and paroxysmal atrial fibrillation for patients in sinus rhythm.^12,13^ Other models have demonstrated predictive power beyond the cardiovascular system, estimating factors such as age, sex, serum potassium, and one-year mortality with a high degree of accuracy.^7,14-16^ However, the full extent of human diseases that may become manifest on the surface ECG remains unknown.

Modern electronic health record (EHR)-based technology has made available large, detailed datasets with rich phenotypic information, enabling large-scale disease-based association testing.^17-19^ Phenome-wide association studies (PheWAS) facilitate high-throughput association testing between predictor variables and multiple disease states using electronically ascertained diagnostic codes. Diseases are commonly represented by phecodes, or standardized, aggregated groupings of International Classification of Disease (ICD) codes.^20-22^

In the present study, we sought to harness both the inferential capabilities of deep learning models and the analytic power of traditional PheWAS to comprehensively assess the array of disease states that manifest in the ECG waveform. Given the growing number of consumer-based wearable devices capable of recording single-lead ECGs, we performed parallel analyses using both traditional 12-lead ECGs and data derived only from lead I, a common single-lead vector used for consumer-based ECG recording.^23^ Specifically, we trained deep learning models known as variational autoencoders to encode 12- and single-lead ECGs within a latent space using a large, unique study sample comprising primary care patients. We selected the autoencoder model because it is unsupervised and optimized to learn ECG waveform features alone, without additional information regarding patient demographics or clinical outcomes. We then used the position of ECG encodings in the latent space to perform high-throughput association testing with roughly 1,600 diseases across three independent datasets spanning over 150,000 individuals. We further demonstrate how latent space modeling can be used to display characteristic ECG features for detectable conditions and to clinically profile individual patients for potentially subclinical diseases that may cause morbidity.

## RESULTS

### Study sample and autoencoder development

Our study utilized three independent datasets spanning over 150,000 individuals, each of which contained individual-level demographic and clinical information, including 12-lead ECGs (**Figure 1**). Two datasets were taken from the Community Care Cohort Project (C3PO), a previously established cohort comprising adults aged ≥ 18 years who received longitudinal primary care within the Mass General Brigham heathcare network between 2000-2018, which is linked by a common EHR data warehouse.^24^ The Massachusetts General Hospital (MGH) C3PO dataset included 60,140 primary care patients and the Brigham and Women’s Hospital (BWH) C3PO dataset included 46,027 primary care patients. The third dataset comprised 35,070 participants from the UK Biobank, a prospective national biorepository that enrolled individuals aged 40-60 years between 2006-2010, with deep phenotyping data, baseline questionnaires, and linkage to electronic health record data.^25^ Characteristics of participants by dataset are provided in **Table 1**.

**Figure 1:**
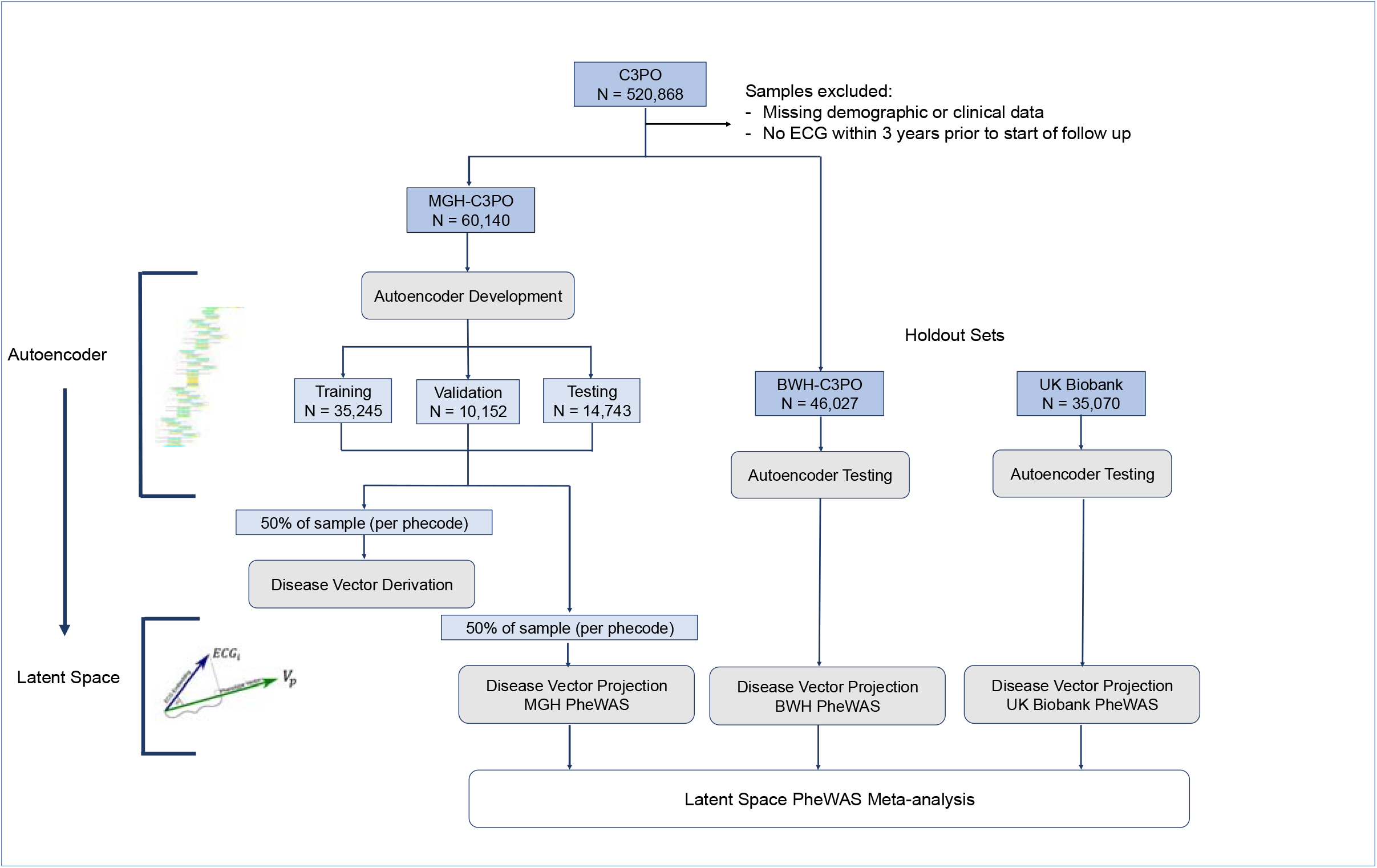
Study overview. Flow diagram of autoencoder and disease vector derivation for latent space phenome-wide association studies (PheWAS), conducted in parallel for both 12-lead and single-lead electrocardiogram (ECG) models. We trained an autoencoder to encode and reconstruct 12- and single-lead ECGs using the Massachusetts General Hospital (MGH) subset of the Community Care Cohort Project (C3PO) dataset (MGH-C3PO). We tested the autoencoder in an MGH-C3PO holdout set and in the independent Brigham and Women’s Hospital (BWH) subset of C3PO (BWH-C3PO), as well as the UK Biobank dataset. We derived disease vectors using labeled ECGs from 50% of the MGH-C3PO dataset. We then projected ECG encodings from the remaining MGH-C3PO samples, as well as from the BWH-C3PO and the UK Biobank datasets, onto these disease vectors. For each ECG encoding, we calculated the projected component, or the position along each disease vector, and tested associations between projected components and corresponding diseases. We performed individual PheWAS in each of the three datasets and then meta-analyzed results.

**Table 1:**
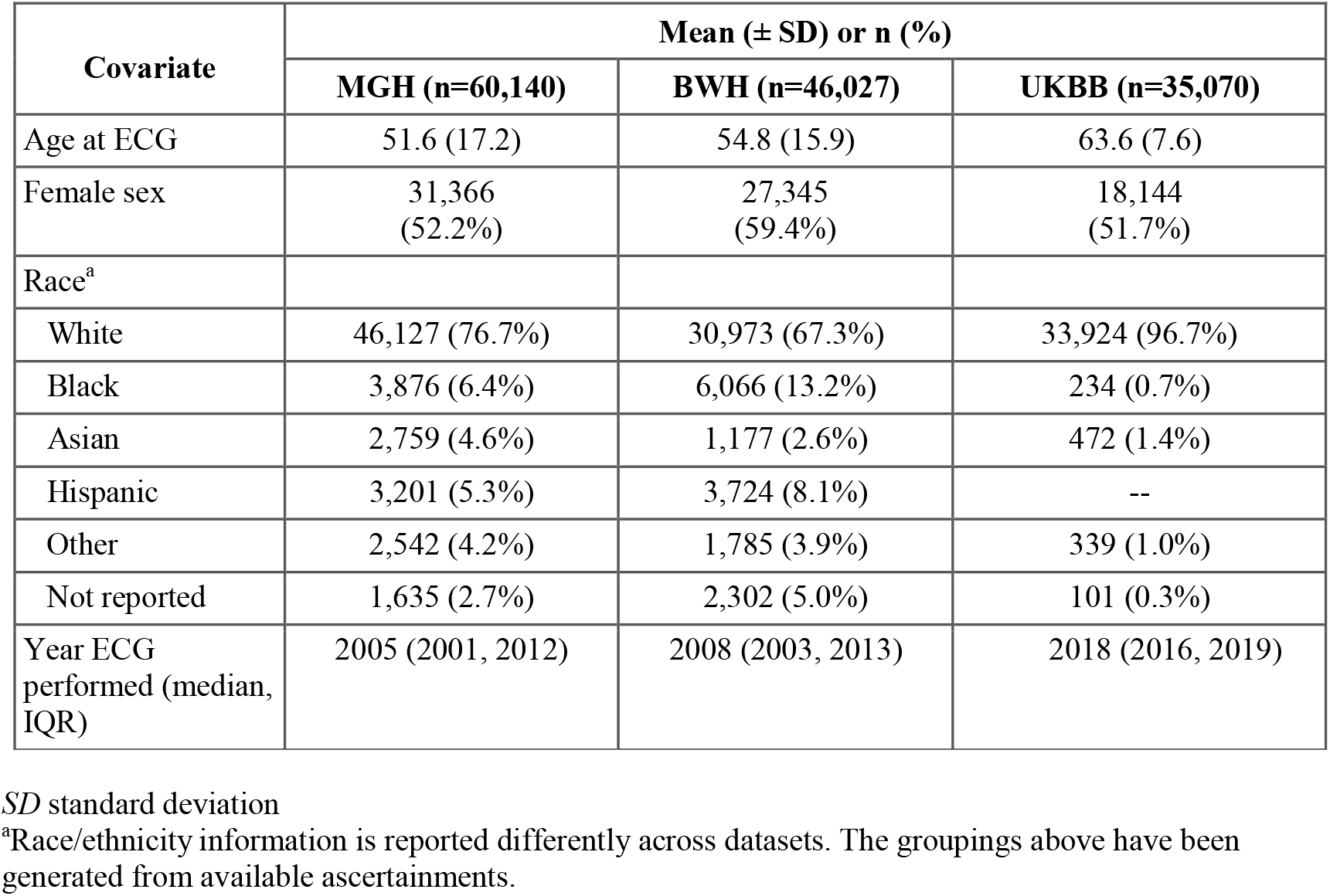
Patient characteristics by dataset.

For both 12- and single-lead models, autoencoders were developed using ECGs from the MGH-C3PO dataset, including 35,245 individuals for training; 10,152 individuals for validation; and 6,674 individuals for testing. The best performing architecture contained over 11 million neurons and used mish activations, 2 dense convolutional blocks (each with 5 layers of convolutions per block), a 71 timestep convolutional kernel, and layer normalization, with 256 neurons in the fully connected layer (**Supplementary Figure 1)**. The models were trained to reconstruct a median waveform corresponding to a single PQRST cycle (**Supplementary Figure 2**). To assess the accuracy of reconstruction in each of the three datasets, we pooled voltages across test set ECGs and compared them to the voltages generated from reconstructions. For each dataset, we assessed the average per-voltage Pearson correlation coefficient and the 95% confidence interval based on 1,000 bootstrap resamplings. We selected this method, as opposed to comparing per-person pooled voltages across test set and reconstructed ECGs, to assess the model’s global performance in ECG reconstruction rather than its performance reproducing ECGs from specific individuals within the datasets. We observed high-fidelity reconstruction of novel ECG median waveforms with Pearson correlation coefficients of 0.9956, 95% CI 0.9931 - 0.9972 in the MGH-C3PO testing set; 0.9916, 95% CI 0.9853 - 0.9950 in BWH-C3PO; and 0.9526, 95% CI 0.9427 - 0.9617 in the UK Biobank.

### Disease vector derivation and ECG projection

For disease vector derivation, we mapped 50% of ECGs from the MGH-C3PO dataset to a library of 1,866 phecodes, grouped across 17 disease categories (e.g., circulatory system, endocrine/metabolic, genitourinary).^21^ Labeled ECG encodings were used to derive disease vectors within the latent space. We then encoded the remaining unlabeled ECGs across the three datasets (n=30,070 for MGH-C3PO; n=46,027 for BWH-C3PO; and n=35,070 for UK Biobank) and determined the position of each ECG along each disease vector. A schematic summarizing the disease vector concept is displayed in **Supplementary Figure 3**.

### Latent space PheWAS

A PheWAS was then performed in the 50% of MGH-C3PO samples that were held-out from disease vector derivation. To assess the validity of results prior to downstream analysis, we performed empiric perturbation testing by randomly reclassifying samples based on disease presence or absence. We generated quantile-quantile plots for MGH-C3PO 12-lead results as well as random disease reclassification levels of 10%, 20%, and 100%. **Supplementary Figure 4** demonstrates that, as expected, the number of significant associations decreases as the degree of random reclassification increases.

PheWAS was additionally performed in the external replication sets and then results from all three PheWas (i.e., the held-out 50% of MGH-C3PO, BWH-C3PO, and UK Biobank) were meta-analyzed to provide a single pooled association test result for each disease. After filtering out diseases with less than 100 combined cases, those present in only one dataset, and those for which model convergence failed, we meta-analyzed the study-specific results for 1,595 diseases for the 12-lead model and 1,600 diseases for the single-lead model (**Figure 2, Supplementary Tables 1 and 2**). Using a Bonferroni-corrected two-sided p-value of 3.1 × 10^-5^ (0.05/1,595 and 0.05/1,600), we observed significant associations between latent space position and disease status for 645 diseases in the 12-lead model (40% of all diseases tested) and 565 diseases in the single-lead model (35%), respectively. Greatest enrichment for associations was observed for diseases of the circulatory (n=140, or 82% of diseases in this category for the 12-lead model, and n=139, or 81% for the single-lead model), respiratory (n=53, 62% category-specific diseases for the 12-lead model; n=46, 54% for the single-lead model) and endocrine/metabolic systems (n=73, 45% of category-specific diseases for the 12-lead model; n=72, 44% for the single lead model; **Figure 2**).

**Figure 2:**
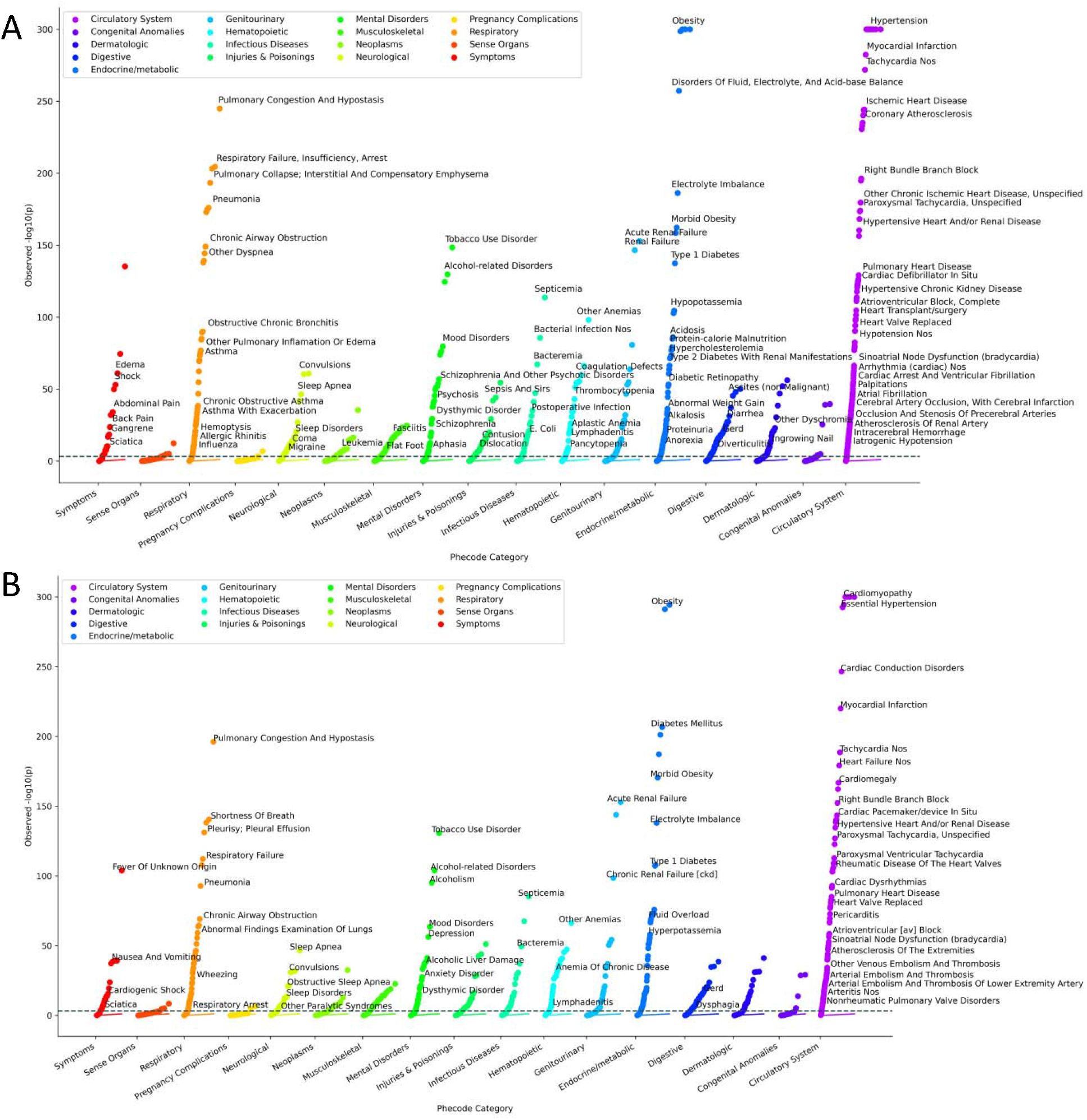
Latent space phenome-wide association study results for 12-lead and 1-lead electrocardiogram autoencoder models. Panel A depicts results for the 12-lead electrocardiogram autoencoder phenome-wide association study, and panel B depicts results for the 1-lead electrocardiogram autoencoder. Each disease tested for association is represented as a single point on the plot. The x-axis represents the disease, or phecode, category and the y-axis represents the -log10(p-value) for the association test. For visual clarity, when disease labels overlap only the more significant one is shown, for the full list of significant diseases see **Supplementary Tables 1 and 2**.

For the 12-lead model, the latent space positions for the phecodes for hypertension, including *hypertension* and *essential hypertension*, showed the strongest associations (p < 2.2 × 10^-308^ for both diseases), followed by *cardiomyopathy*(p < 2.2 × 10^-308^). The strongest associations among non-cardiac diseases included *obesity* (p < 2.2 × 10^-308^), *diabetes mellitus* (p = 5.9 ×10^-304^), *disorders of fluid, electrolyte, and acid-base balance* (p = 5.5 × 10^-258^), and *pulmonary congestion and hypostasis* (p = 1.2 × 10^-245^). For the single-lead model, the latent space positions for the phecodes for cardiomyopathy, including *cardiomyopathy* and *primary/intrinsic cardiomyopathies*, showed the strongest associations (p < 2.2 × 10^-308^ for both), followed by *congestive heart failure NOS* (p < 2.2 × 10^-308^). Forest plots summarizing the associations for the top diseases are displayed in **Supplementary Figure 5**.

We additionally identified unexpected and highly robust relationships, including tobacco use disorder (p = 1.0 × 10^-149^ for 12-lead and p = 1.9 × 10^-131^ for single-lead), fever of unknown origin (p = 5.5 × 10^-136^ and 1.2 × 10^-104^), and non-alcoholic liver disease (p = 8.0 × 10^-51^ and 1.9 × 10^-39^). Importantly, effects were generally consistent across datasets, including in the UK Biobank, which had an overall lower prevalence of disease (**Supplementary Tables 1 and 2**).

### ECG intervals PheWAS

Meta-analyses of the ECG intervals PheWAS included 1,607 diseases for the PR interval; 1,607 diseases for the QRS duration; and 1,605 diseases for the QT interval. We compared the ECG intervals and latent space models by restricting to diseases that were present in all meta-analyses (n=1,584). For the ECG interval analyses, we took the smallest p-value for each disease across each of the ECG intervals tested. We observed the greatest absolute number of significant associations in the 12-lead latent space model, followed by the single-lead latent space model, and the fewest in the ECG intervals model, both overall and within disease categories (**Figure 3** and **Supplementary Tables 3-5**).

**Figure 3:**
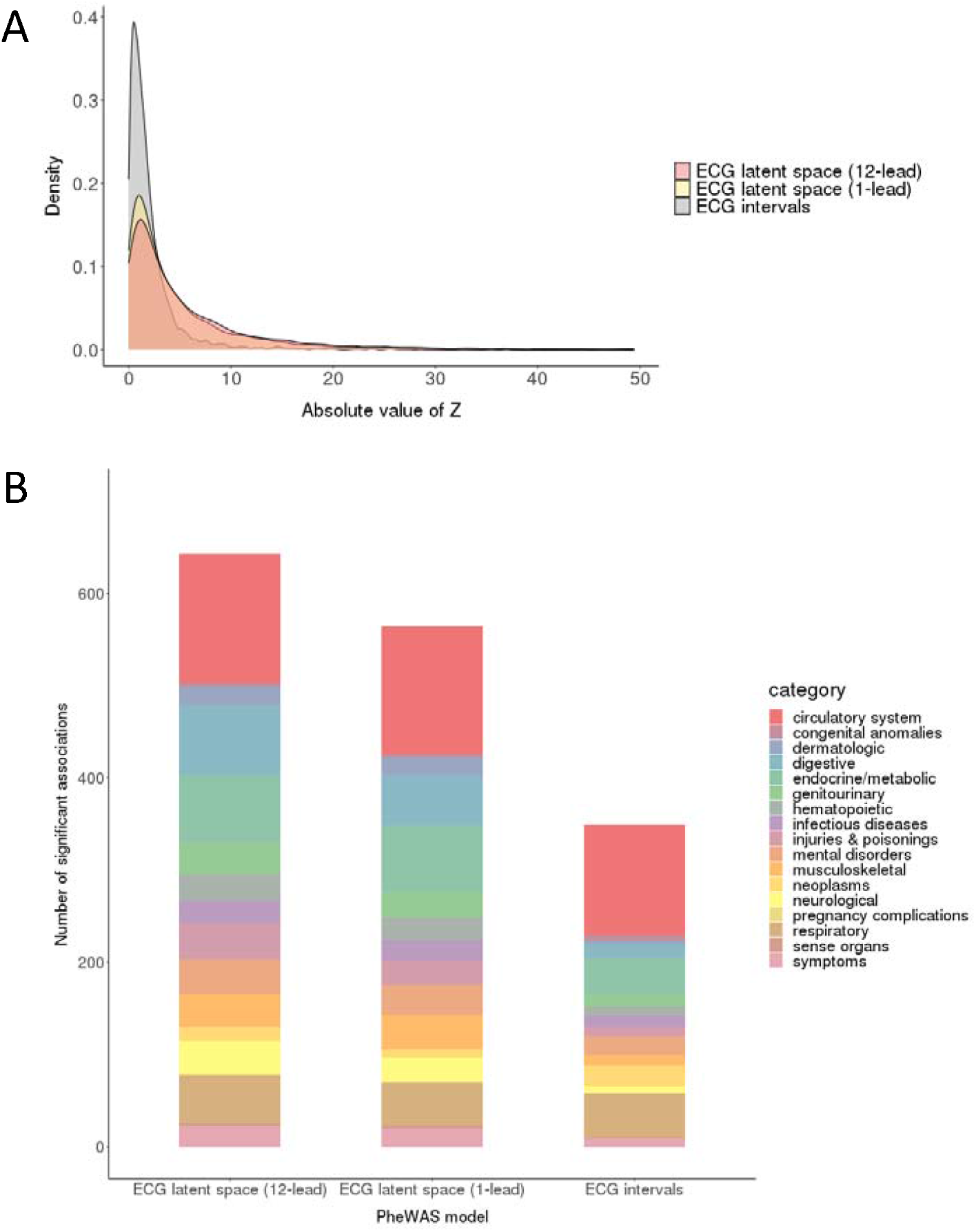
Significant associations in the latent space versus electrocardiogram intervals phenome-wide association studies. Panel A displays the test statistic distribution (absolute z-score) for the ECG term in the meta-analyzed phenome-wide association study (PheWAS), stratified by modeling approach. Results are displayed for the 12-lead and 1-lead electrocardiogram (ECG) latent space models, as well as the ECG intervals model. Panel B demonstrates the number of significantly associated diseases, defined as those exceeding a Bonferroni-corrected two-sided p-value of 3.2 × 10^-5^ (0.05 divided by 1,584, the number of unique diseases included across all meta-analyses). For the intervals model, a result was considered significant if the meta-analyzed p-value for any of the tested ECG intervals (PR, QRS, QT) exceeded the significance threshold. When compared to the ECG intervals model, the latent space models yield a greater number of significant associations, both overall and across disease categories.

### Discrimination of diseases

We then compared discrimination of diseases using the area under the receiver operating characteristic curve (AUC) across logistic regression models in which diseases were regressed on age, sex, and race with or without an additional term for projected ECG component. We restricted analyses of discrimination to those diseases for which a significant difference in the ECG encoding was observed along the disease vector from the PheWAS analysis (p<3.1×10^-5^). We generally observed a substantial increase in discrimination for models that included the ECG latent space projected components (see Methods for derivation) as compared to models that did not. For example, improvements in the AUC were observed for the circulatory system (median difference in AUC, interquartile range for MGH-C3PO 0.034, 0.019-0.069; BWH-C3PO 0.027, 0.011-0.062; UKB 0.005, 0.001-0.014) and respiratory (MGH-C3PO 0.044, 0.024-0.073; BWH-C3PO 0.027, 0.011-0.059; UKB 0.002, 0.000-0.005) disease categories, with improvements in discrimination observed for other disease categories as well (**Supplementary Figure 6**).

### Model-Based, disease-specific median waveforms

For certain diseases with well-characterized ECG manifestations, model-derived features were consistent with expectation. For example, median waveforms generated from the left bundle branch block disease-positive centroid demonstrated QRS widening, smaller initial r waves in the right-sided precordial leads (V1-V3), and R wave slurring in the left-sided leads (I, aVL, V5, V6)^26^ relative to median waveforms generated from the disease-negative centroid (**Figure 4A**). For hypokalemia, model-derived features included decreased T wave amplitude and relative QT prolongation^27^ (**Figure 4B**). In other instances, however, reconstructed disease case and control ECGs appeared morphologically similar despite highly significant differences in latent space position detected by PheWAS. For example, model-derived ECG reconstructions of hypertrophic cardiomyopathy were notable for broad, flattened T-waves, particularly in the left-sided leads, which are more subtle than the classical well-defined ECG manifestations of hypertrophic cardiomyopathy (e.g., prominent precordial voltages, repolarization abnormalities/T-wave inversions, pathologic Q waves^28^) (**Figure 4C**). The findings suggest that the latent space is sensitive to subtle waveform manifestations of disease, while the model-derived ECG reconstructions are conservative and may not visually replicate all hallmarks of disease.

**Figure 4:**
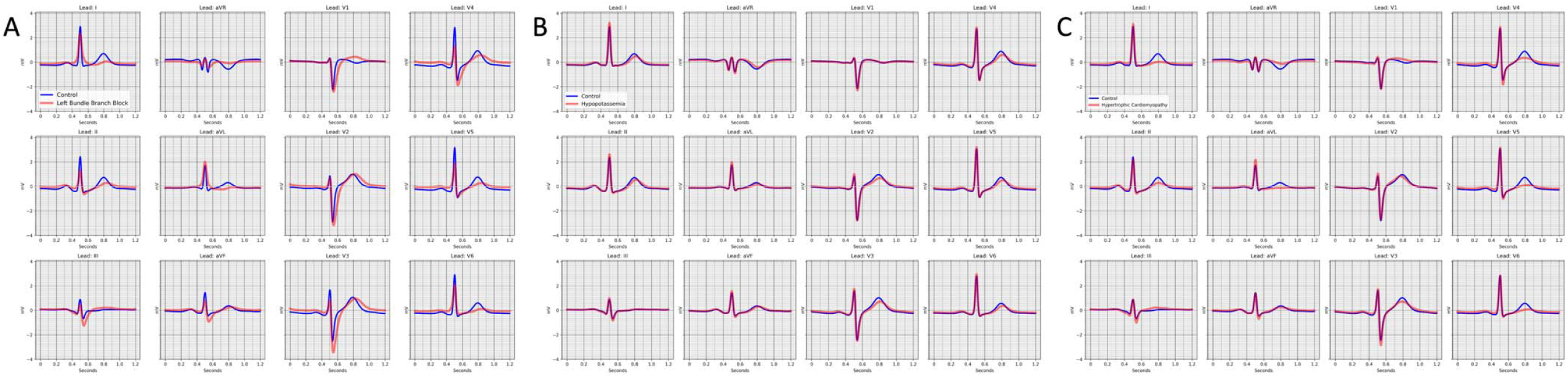
Model-based, disease-specific ECG reconstructions. Median wave-form reconstructions for centroids reflecting individuals without (blue) and with (red) Left bundle branch block in panel A, hypopotassemia (hypoptassemia) in panel B, and Hypertrophic obstructive cardiomyopathy in panel C.

### Patient report card prototype

To further demonstrate the potential for the ECG to serve as a digital biomarker for disease status, we plotted a prototype of a patient report for select diseases (**Table 2**). In this illustrative example, an ECG from a 65-year-old female is projected onto the disease vectors for select circulatory system diseases, and the positions relative to the whole cohort along vectors from the disease negative to disease positive centroids are reported. These diseases were selected based on clinical relevance and the potential to cause substantial morbidity if undetected, including stroke, sudden cardiac death, and heart failure.^29-33^

**Table 2.** Patient report prototype for a 65-year-old female. A representative example of a patient report for one randomly selected individual. Disease proximity is expressed as the percent distance along a normalized disease vector, from the disease negative to the disease positive centroids. The report highlights diseases that were selected for illustrative purposes based on statistical significance as well as clinical relevance. The expected value for an average risk in the dataset along the normalized disease vector is 50% given that each disease vector was normalized to have length one, and the entire space of ECG encodings was scaled to have norm and standard deviation of one.

| Disease | Distance along normalized disease vector |
| --- | --- |
| Hypertrophic cardiomyopathy | 48% |
| Atrial fibrillation | 71% |
| Cardiomyopathy | 46% |
| Heart failure with preserved ejection fraction | 33% |
| Hypertension | 38% |
| Myocardial infarction | 43% |
| Paroxysmal ventricular tachycardia | 56% |
| Mitral and aortic valve stenosis | 52% |

## DISCUSSION

Here, we highlight a novel application of autoencoder deep learning models, specifically demonstrating how a model trained to encode and reconstruct 12- and single-lead ECGs generates a multidimensional latent space that can be used to explore relations between ECG waveform features and clinically relevant disease phenotypes at scale. Our results suggest robust associations between ECG waveform patterns and diseases across multiple organ systems. When compared to an ECG intervals model, autoencoder-based latent space models reveal a greater number of significant associations, suggesting that the architectural complexity provided by the deep learning model is accompanied by substantially greater information about disease status than routinely ascertained intervals.

Autoencoder models have been used previously in fields such as linguistics and image processing to better understand input information. Prior studies on image representation have demonstrated that attribute labeling (e.g., headshot photograph categorized as *wearing glasses* versus *no glasses*) can help understand the machine-learned latent space, as images with similar attributes form interpretable semantic clusters. Cluster centroids are spatially defined using attribute vectors, and any unlabeled image projection can be relationally defined by its position along the vector, based solely on the image characteristics interpreted by the model. These studies further demonstrate that the reconstruction of an input image (e.g., person without glasses) can be modified by movement along an attribute vector, generating a novel image with features that are more aligned with the corresponding centroid pair (e.g., same photograph reconstructed with glasses).^34,35^

Our study applies autoencoder-based technology to clinical ECG data, yielding results with several implications. First, we demonstrate how latent space modeling can be used for the discovery of novel information contained within a widely available and inexpensive diagnostic modality, the ECG. Specifically, we apply attribute, or in this case disease, labelling to explore disease-state information contained in 12- and single-lead ECGs. By reconstructing median waveforms from latent space centroids, we visually represent disease-based patterns identified by deep learning models, which in some cases confirm expectations and in other cases reflect subtle waveform manifestations that may not be discernible by human interpretation. Direct extensions of ECG-based latent space modeling could provide insight into the prediction of incident disease, or the natural history of existing disease, using samples acquired prior to and after a given diagnosis for derivation of “disease progression vectors,” allowing for the direct visualization of waveform evolution over time. We submit that the methodology of mapping clinical status onto the latent space may have implications far beyond the ECG, extending to other modalities (e.g., laboratory testing, imaging results) and/or multimodal testing, thereby greatly expanding the clinical utility of existing, easily acquired diagnostics.

Second, while ECG-based deep learning models have been developed previously, studies have predominantly focused on disease-specific risk prediction within the cardiovascular system.^8,9,12,13^ In taking a more global analytic approach, we have identified a potential role for ECG-based classification of non-cardiac disease. We additionally highlight conditions for which further study may be particularly high yield, including diseases not classically associated with ECG findings but each independently supported by prior studies (e.g., type 2 diabetes,^36,37^ sleep apnea,^38-41^ chronic liver disease/cirrhosis,^15,42^ and renal failure^43^), as well as diseases with previously undescribed associations.

Third, we demonstrate the potential for personalized and scalable disease detection with ECG-based latent space modeling. Using data derived from large samples, we construct a complex architectural environment informed by disease status, in which each ECG encoding represents a single individual and occupies a unique position in the latent space. Projection of new ECGs from independent individuals can therefore be used to generate likelihoods of disease at scale. As latent space modeling approaches are refined with data from larger and more diverse samples, we anticipate that the utility for disease-based classification will grow. Our patient report card is illustrative in nature, and future studies are warranted to evaluate the specific test characteristics of latent space proximity in discriminating disease status. We submit that the approach we outline will have particular value for detecting diseases in which screening may be cumbersome, inaccurate, or expensive and for which early disease manifestations may be highly morbid (e.g., hypertrophic cardiomyopathy and dilated cardiomyopathy). Indeed, the robust performance of our single-lead model for prevalent disease detection highlights the scalable utility for screening large populations, particularly given the widespread emergence of consumer-based wearable and handheld devices with ECG recording capabilities.^23^

Our study presents certain limitations. As discussed, disease states were represented by diseases generated from prevalent ICD codes. For this reason, our models are trained to highlight relations between the ECG and existing diseases and are otherwise not optimized to identify ECG features that might predict incident disease. Additionally, diagnosis codes may be inaccurate and/or temporally unrelated to the ECG sampled, particularly in instances of acute or paroxysmal disease, leading to misclassification bias. While we suggest the high signal intensity within the circulatory system category provides evidence of internal validity, we cannot exclude the possibility of confounding by indication, in which diseases of the circulatory system are over-represented because ECGs are more likely to be performed in cases where cardiac disease is suspected. However, our findings were generally consistent in the UK Biobank, in which ECGs were obtained systematically and independent of clinical indications. Finally, because the UK Biobank represents a relatively healthy population, PheWAS in this dataset is likely underpowered, thereby producing false negative results within our meta-analysis.

In conclusion, we demonstrate the potential utility of latent space modeling to extract clinically relevant information embedded within currently available diagnostics. Our findings suggest that ECG waveform data contain a wealth of disease-state information extending far beyond the circulatory system. Future studies should further evaluate the significant potential of 12- and single-lead waveform analysis for scalable disease profiling.

## Data Availability

The Mass General Brigham source data are not publicly available because they are electronic health records. Making the data publicly available without additional consent or ethical approval could compromise privacy. Source data from the UK Biobank are available to qualified investigators via application at https://www.ukbiobank.ac.uk.

## METHODS

### Study subjects

Two of the three datasets included were derived from the Community Care Cohort Project (C3PO), a previously established cohort comprising over 500,000 adults aged ≥ 18 years who receive longitudinal primary care at one of eleven hospitals within the Mass General Brigham network, which is linked by a common EHR data warehouse.^44^ C3PO datasets included a cohort from the Massachusetts General Hospital (MGH-C3PO dataset) and a cohort from the Brigham and Women’s Hospital (BWH-C3PO dataset). The third, external dataset was derived from the UK Biobank, a prospective community-based cohort study comprising adults aged 40-60 years at enrollment between the years 2006-2010 from the United Kingdom.^25^ The present analysis includes the subsets of individuals in each dataset with at least one 12-lead ECG performed within three years prior to the start of follow up (C3PO) or who had a 12-lead ECG performed during at least one study visit (UK Biobank). Use of MGB and UK Biobank (application 7089) data were approved by the MGB Institutional Review Board. The UK Biobank was approved by the UK Biobank Research Ethics Committee (reference number 11/NW/0382). All UK Biobank participants provided written informed consent.

### ECG autoencoder model and latent space derivation

We trained densely connected convolutional autoencoders to encode and reconstruct 12- and single-lead ECGs. In general, autoencoders consists of an encoder, which maps a high-dimensional input into a lower-dimensional latent space, and a decoder, which reconstructs the original data from the latent space representation (**Supplementary Figure 1**). Autoencoders are trained to encode variance present within the original data into the latent space, which encourages the model to minimize differences between the original data and its reconstruction. Both the 12-lead and single-lead autoencoders were trained and validated using subsets of ECGs from the MGH-C3PO cohort. The models were then tested in an MGH-C3PO holdout set as well as two external holdout datasets, including BWH-C3PO and the UK Biobank.

To standardize the phase of the cardiac cycle across all individuals while minimizing the effects of signal artifact (e.g., baseline drift, transient noise), we encoded ECGs as median waveforms by segmenting 10-second ECG recordings into 1,200 millisecond windows, sampling 600 voltage timepoints per window, and performed piecewise linear interpolation to generate R-R adjusted medians.^45,46^ Median waveforms therefore represent the aggregate morphology of at least one cardiac cycle from each lead (**Supplementary Figure 2**). The 12-lead model utilized median waveforms generated from all available leads (i.e., 12 waveforms per ECG), while the single-lead model utilized only the median waveform generated from lead I. In the following analysis, the term ECG generally refers to the median waveform.

Models were trained using one-dimensional convolutions over voltage-time series, corresponding to 7,200 voltage timepoints for 12-lead ECGs and 600 voltage timepoints for single leads. For ECGs with incomplete voltage data (i.e., less than 10 seconds recorded from each lead), we used zero padding, converting non-available data into zeros (**Supplementary Table 6**). The mean squared error per voltage timepoint across the full ECG was minimized, as demonstrated in equation (1):

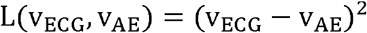

In C3PO, ECGs were excluded if the acquisition date was greater than three years prior to the start of clinical follow up, defined for each individual as the time of the second primary care visit of the earliest qualifying pair.^44^ Only one ECG per individual was represented; for patients with multiple ECGs, the most recent was used.

The neural net architecture was a variant of Densenet, featuring several densely connected convolutional blocks operating at different time resolutions.^47^ Architecture hyperparameters including width, depth, activation, normalization, and regularization were chosen via Bayesian hyperparameter optimization.^48^

To assess the accuracy of reconstruction in each of the three datasets, we pooled voltages across test set ECGs and compared this to the voltages generated from reconstructions. For each dataset, we assessed the average per-voltage Pearson correlation coefficient and the 95% confidence interval based on 1,000 bootstrap resamplings. We selected this method, as opposed to comparing per-person pooled voltages across test set and reconstructed ECGs, to assess the model’s global performance in ECG reconstruction rather than its performance reproducing ECGs from specific individuals within the datasets (**Supplementary Figure 2**).

### Disease definitions

Diseases in each of the three datasets were mapped using ICD codes to a publicly available phecode library (https://phewascatalog.org/phecodes_icd10cm).^20,49^ As previously described, phecodes distinguish cases from controls using hierarchical groupings of ICD 9 and 10 codes to better define clinically meaningful disease phenotypes. Only prevalent phecodes were used, i.e., all corresponding ICD codes had been entered into the patient’s chart prior to the ECG acquisition date. For certain phecodes, participants without that phecode but with very similar ICD codes were excluded from serving as controls to avoid biasing results, as described previously (e.g., in association testing for the myocardial infarction case group, patients were removed from serving as controls if they had ICD codes corresponding to a list of disease exclusions, including angina or other evidence of ischemic heart disease).^20,21^

### Disease vector derivation

If a given disease, represented by a phecode, has a significant impact on the ECG, we expect ECG encodings from individuals with the disease to distribute to a different location in the autoencoder-derived latent space relative to ECG encodings from individuals without the disease. In contrast, if the disease has little impact on the ECG, or if the ECG encoding does not adequately capture disease-relevant features, then we expect there to be no significant relationship between the position of the ECG encoding in latent space and the presence or absence of disease.

To quantify this expectation, we define the highest density of ECG encodings labeled as having the disease (“disease-positive centroid” for cases) and the highest density of ECG encodings labeled as not having the disease (“disease-negative centroid” for controls). Each disease is therefore spatially represented by its centroid pair, and the line that connects them is referred to as the disease vector (**Supplementary Figure 3**).

During autoencoder training/latent space derivation, we labeled a subset of ECGs from the MGH-C3PO dataset as disease cases or controls based on the presence or absence of the corresponding disease in the patient’s EHR. ECG encodings from the derivation set were then used to define centroid pairs and disease vectors for each disease within the latent space. The disease vector correlation matrix is displayed in **Supplementary Figure 7**.

### ECG projections

Any ECG encoded within the latent space, including encodings from unlabeled samples, can be projected onto any disease vector, and the relative position along the disease vector can be used to assess how closely related the unlabeled ECG encoding is to encodings that were used to define the disease positive centroid for that disease.

After deriving disease vectors in our sample, we projected unlabeled ECG encodings from the MGH-C3PO holdout set, as well as from our two external datasets, BWH-C3PO and the UK Biobank, onto each disease vector generated within the latent space. We scaled the entire space of ECG encodings to have norm and standard deviation of one. Each disease vector was normalized to have length one. The high-dimensional spatial relatedness of each ECG and disease was quantified by each sample’s component in the direction of a given disease vector. As illustrated below in equation (2), each ECG encoding (“ECG embedding”, ECG_i_) projects onto each disease vector, V_p_. The projected component, (“component_ip_”) is calculated from the angle between the ECG encoding and the normalized disease vector. Thus, the projected component signifies the latent space position of a single individual along a single disease vector.

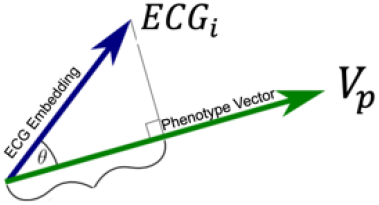

We used analogous methods for both 12- and single-lead models. In the single-lead model, the autoencoder-derived latent space, disease vector derivation, and ECG projections were based only on median waveform data derived from lead I.

### Association Testing by Latent Space PheWAS

For both 12- and single-lead models, we performed a PheWAS in each dataset using a logistic regression model to assess the strength of the relationship between the position of a latent space ECG encoding (using the projected component as the predictor variable of interest) and a given disease state (using disease presence or absence as the outcome variable). For the UK Biobank dataset, the model was adjusted for age, sex, and race. For the MGH-C3PO holdout set and the BWH-C3PO dataset, the model was additionally adjusted for ECG acquisition date and the amount of zero padding, which qunaitified the degree of missing voltage data and may represent other unmeasured confounders such as ECG quality, ECG machine used, hospital location, etc. Discrimination was assessed using the area under the receiver operating characteristic curve.

For both latent space models, we performed a fixed effects inverse variance weighted meta-analysis, filtering for diseases that were present in at least two datasets with at least 100 combined cases. Coefficients corresponding to each disease vector were pooled across datasets. For comparison, we generated a separate model based on ECG intervals, including the PR interval, QRS duration, and QT interval. We chose an intervals-based model as our comparator because it utilizes routinely ascertained, standardized, and automated measurements known to have disease-based prognostic implications.^50-54^ As above, we performed an intervals PheWAS in each dataset and meta-analyzed results. We then compared the number of significant associations between the latent space models and the ECG intervals model, using a Bonferroni-corrected two-sided p-value of 0.05 divided by the number of common diseases across all meta-analyses. For the intervals model, we considered a result significant if the p-value for any interval met the significance threshold. Metanalyses were performed using the R package *meta*. Python was used to create a visual summary of meta-analyzed data, grouped according to disease category.

### Disease-based ECG reconstructions

To better understand which ECG features may have contributed to disease segregation in latent space, we generated disease-specific median waveforms. Specifically, we decoded ECGs from disease-positive centroids and overlayed the resultant median waveforms on ECGs decoded from the corresponding disease-negative centroids.

### Patient report card

One advantage of latent space modeling is the ability to incorporate an enormous amount of ECG- and EHR-based data in a multidimensional environment. The multidimensional nature of the model allows for simultaneous assessment of an ECG encoding’s proximity to all disease centroids. We sought to demonstrate the associated potential for personalized and scalable disease reporting by converting the position of an ECG encoding for a single individual into a percent distance along normalized disease vectors from the disease negative (0%) to the disease positive (100%) centroids.

## CODE AVAILABILITY

The JEDI data processing pipeline underlying C3PO, as well as data processing scripts underlying the current analyses, are available in our opensource github repository at https://github.com/broadinstitute/ml4h.

## ACKNOWLEDGMENTS

Dr. Lubitz is a full-time employee of Novartis Institutes for Biomedical Research as of July 18, 2022. Dr. Lubitz previously received support from NIH grants R01HL139731 and R01HL157635, and American Heart Association 18SFRN34250007. Dr. Anderson is supported by NIH grants R01NS103924 and U01NS069763 and American Heart Association grants 18SFRN34250007 and 21SFRN812095. Dr. Weng is supported by National Institutes of Health (NIH) grant 1R01HL139731. Dr. Choi is supported by the NHLBI BioData Catalyst Fellows program. Dr. Ellinor is supported by the NIH (1R01HL092577, K24HL105780), AHA (18SFRN34110082) and by MAESTRIA (965286). Dr. Lau is supported by the American Heart Association (853922).

## REFERENCES

1. Trobec, R. & Tomašić, I. Synthesis of the 12-lead electrocardiogram from differential leads. IEEE Trans Inf Technol Biomed 15, 615–621 (2011).

2. Barold, S.S. Willem Einthoven and the birth of clinical electrocardiography a hundred years ago. Card Electrophysiol Rev 7, 99–104 (2003).

3. Rivera-Ruiz, M., Cajavilca, C. & Varon, J. Einthoven’s string galvanometer: the first electrocardiograph. Tex Heart Inst J 35, 174–178 (2008).

4. Salvati, M., et al. Electrocardiographic changes in subarachnoid hemorrhage secondary to cerebral aneurysm. Report of 70 cases. Ital J Neurol Sci 13, 409–413 (1992).

5. Surawicz, B. Relationship between electrocardiogram and electrolytes. Am Heart J 73, 814–834 (1967).

6. Van Mieghem, C., Sabbe, M. & Knockaert, D. The clinical value of the ECG in noncardiac conditions. Chest 125, 1561–1576 (2004).

7. Yasin, O.Z., et al. Noninvasive blood potassium measurement using signal-processed, single-lead ecg acquired from a handheld smartphone. J Electrocardiol 50, 620–625 (2017).

8. Al-Zaiti, S., et al. Machine learning-based prediction of acute coronary syndrome using only the pre-hospital 12-lead electrocardiogram. Nat Commun 11, 3966 (2020).

9. Cikes, M., et al. Machine learning-based phenogrouping in heart failure to identify responders to cardiac resynchronization therapy. Eur J Heart Fail 21, 74–85 (2019).

10. Feeny, A.K., et al. Artificial Intelligence and Machine Learning in Arrhythmias and Cardiac Electrophysiology. Circ Arrhythm Electrophysiol 13, e007952 (2020).

11. Hannun, A.Y., et al. Cardiologist-level arrhythmia detection and classification in ambulatory electrocardiograms using a deep neural network. Nat Med 25, 65–69 (2019).

12. Attia, Z.I., et al. Screening for cardiac contractile dysfunction using an artificial intelligence-enabled electrocardiogram. Nat Med 25, 70–74 (2019).

13. Attia, Z.I., et al. An artificial intelligence-enabled ECG algorithm for the identification of patients with atrial fibrillation during sinus rhythm: a retrospective analysis of outcome prediction. Lancet 394, 861–867 (2019).

14. Attia, Z.I., et al. Age and Sex Estimation Using Artificial Intelligence From Standard 12-Lead ECGs. Circ Arrhythm Electrophysiol 12, e007284 (2019).

15. Galloway, C.D., et al. Development and Validation of a Deep-Learning Model to Screen for Hyperkalemia From the Electrocardiogram. JAMA Cardiol 4, 428–436 (2019).

16. Raghunath, S., et al. Prediction of mortality from 12-lead electrocardiogram voltage data using a deep neural network. Nat Med 26, 886–891 (2020).

17. Barak-Corren, Y., et al. Predicting Suicidal Behavior From Longitudinal Electronic Health Records. Am J Psychiatry 174, 154–162 (2017).

18. Liu, C., Wang, F., Hu, J. & Xiong, H. Temporal Phenotyping from Longitudinal Electronic Health Records: A Graph Based Framework. in Proceedings of the 21th ACM SIGKDD International Conference on Knowledge Discovery and Data Mining 705–714 (Association for Computing Machinery, Sydney, NSW, Australia, 2015).

19. Zhao, J., et al. Learning from Longitudinal Data in Electronic Health Record and Genetic Data to Improve Cardiovascular Event Prediction. Sci Rep 9, 717 (2019).

20. Denny, J.C., et al. Systematic comparison of phenome-wide association study of electronic medical record data and genome-wide association study data. Nat Biotechnol 31, 1102–1110 (2013).

21. Denny, J.C., et al. PheWAS: demonstrating the feasibility of a phenome-wide scan to discover gene-disease associations. Bioinformatics 26, 1205–1210 (2010).

22. Gao, M., Quan, Y., Zhou, X.H. & Zhang, H.Y. PheWAS-Based Systems Genetics Methods for Anti-Breast Cancer Drug Discovery. Genes (Basel) 10(2019).

23. Al-Alusi, M.A., Ding, E., McManus, D.D. & Lubitz, S.A. Wearing Your Heart on Your Sleeve: the Future of Cardiac Rhythm Monitoring. Curr Cardiol Rep 21, 158 (2019).

24. Khurshid, S., et al. Cohort design and natural language processing to reduce bias in electronic health records research. NPJ Digit Med 5, 47 (2022).

25. Littlejohns, T.J., Sudlow, C., Allen, N.E. & Collins, R. UK Biobank: opportunities for cardiovascular research. European Heart Journal 40, 1158–1166 (2017).

26. Tan, N.Y., Witt, C.M., Oh, J.K. & Cha, Y.M. Left Bundle Branch Block: Current and Future Perspectives. Circ Arrhythm Electrophysiol 13, e008239 (2020).

27. Levis, J.T. ECG diagnosis: hypokalemia. Perm J 16, 57 (2012).

28. Finocchiaro, G., et al. The electrocardiogram in the diagnosis and management of patients with hypertrophic cardiomyopathy. Heart Rhythm 17, 142–151 (2020).

29. Wolf, P.A., Abbott, R.D. & Kannel, W.B. Atrial fibrillation as an independent risk factor for stroke: the Framingham Study. Stroke 22, 983–988 (1991).

30. Velagaleti, R.S., et al. Long-term trends in the incidence of heart failure after myocardial infarction. Circulation 118, 2057–2062 (2008).

31. Maron, B.J., et al. Efficacy of implantable cardioverter-defibrillators for the prevention of sudden death in patients with hypertrophic cardiomyopathy. N Engl J Med 342, 365–373 (2000).

32. Wang, T.J., et al. Temporal relations of atrial fibrillation and congestive heart failure and their joint influence on mortality: the Framingham Heart Study. Circulation 107, 2920–2925 (2003).

33. Tsao, C.W., et al. Heart Disease and Stroke Statistics-2022 Update: A Report From the American Heart Association. Circulation 145, e153–e639 (2022).

34. Liu, Y., Jun, E., Li, Q. & Heer, J. Latent Space Cartography: Visual Analysis of Vector Space Embeddings. Computer Graphics Forum (Proc. EuroVis) (2019).

35. Xiao, L. & Wang, J. LatentVis: Investigating and Comparing Variational Auto-Encoders via Their Latent Space. in 3rd Workshop Advances in Interpretable Machine Learning and Artificial Intelligence (AIMLAI) of CIKM (2020).

36. Cordeiro, R., Karimian, N. & Park, Y. Hyperglycemia Identification Using ECG in Deep Learning Era. Sensors (Basel) 21(2021).

37. Wang, L., Mu, Y., Zhao, J., Wang, X. & Che, H. IGRNet: A Deep Learning Model for Non-Invasive, Real-Time Diagnosis of Prediabetes through Electrocardiograms. Sensors (Basel) 20(2020).

38. Li, A., Chen, S., Quan, S.F., Powers, L.S. & Roveda, J.M. A deep learning-based algorithm for detection of cortical arousal during sleep. Sleep 43(2020).

39. Mukherjee, D., Dhar, K., Schwenker, F. & Sarkar, R. Ensemble of Deep Learning Models for Sleep Apnea Detection: An Experimental Study. Sensors (Basel) 21(2021).

40. Urtnasan, E., Park, J.U., Joo, E.Y. & Lee, K.J. Identification of Sleep Apnea Severity Based on Deep Learning from a Short-term Normal ECG. J Korean Med Sci 35, e399 (2020).

41. Sun, H., et al. Sleep staging from electrocardiography and respiration with deep learning. Sleep 43(2020).

42. Toma, L., et al. Electrocardiographic Changes in Liver Cirrhosis-Clues for Cirrhotic Cardiomyopathy. Medicina (Kaunas) 56(2020).

43. Deo, R., et al. Electrocardiographic Measures and Prediction of Cardiovascular and Noncardiovascular Death in CKD. J Am Soc Nephrol 27, 559–569 (2016).

44. Khurshid, S., et al. Cohort Design and Natural Language Processing to Reduce Bias in Electronic Health Records Research: The Community Care Cohort Project. medRxiv, 2021.2005.2026.21257872 (2021).

45. Carreiras, C., et al. Biosppy: Biosignal processing in python. (2015).

46. Verweij, N., et al. The Genetic Makeup of the Electrocardiogram. Cell Syst 11, 229-238.e225 (2020).

47. Iandola, F.N., et al. DenseNet: Implementing Efficient ConvNet Descriptor Pyramids. CoRR abs/1404.1869(2014).

48. Snoek, J., Larochelle, H. & Adams, R.P. Practical Bayesian optimization of machine learning algorithms. in Proceedings of the 25th International Conference on Neural Information Processing Systems- Volume 2 2951–2959 (Curran Associates Inc., Lake Tahoe, Nevada, 2012).

49. Denny, J.C., et al. Phecode Map 1.2 with ICD-10cm Codes (beta). Vol. 2022.

50. Brenyo, A. & Zaręba, W. Prognostic significance of QRS duration and morphology. Cardiol J 18, 8–17 (2011).

51. Castiglione, A. & Odening, K. [QT Interval and Its Prolongation - What Does It Mean?]. Dtsch Med Wochenschr 145, 536–542 (2020).

52. Chen, X., et al. Incremental changes in QRS duration as predictor for cardiovascular disease: a 21-year follow-up of a randomly selected general population. Sci Rep 11, 13652 (2021).

53. Estrada, A.H., et al. Diagnostic accuracy of computer aided electrocardiogram analysis in dogs. J Small Anim Pract 62, 145–149 (2021).

54. Rasmussen, P.V., et al. Electrocardiographic PR Interval Duration and Cardiovascular Risk: Results From the Copenhagen ECG Study. Can J Cardiol 33, 674–681 (2017).

