## Supplementary Materials for "Deep Learning of Electrocardiograms Enables Scalable Human Disease Profiling"

### **Supplementary Figure 1.** General autoencoder schematic.

General autoencoder structure. The input, X, is encoded into a multi-dimensional latent space, h. This “code” is then used to reconstruct the input as accurately as possible at the output layer, X’.

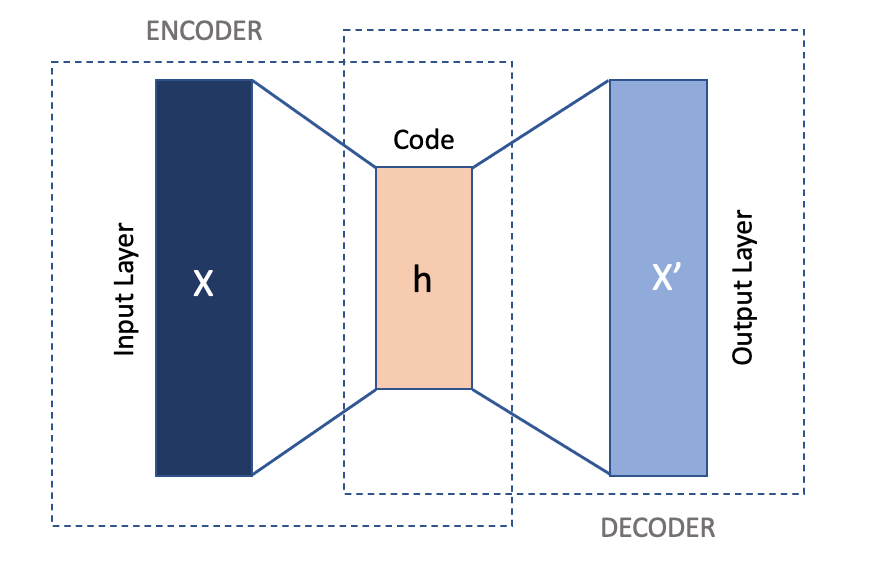

### **Supplementary Figure 2.** Example of median waveforms generated from an original 12-lead electrocardiogram.

Panel A represents the original 12-lead tracing, and panel B represents the median waveform for each lead.

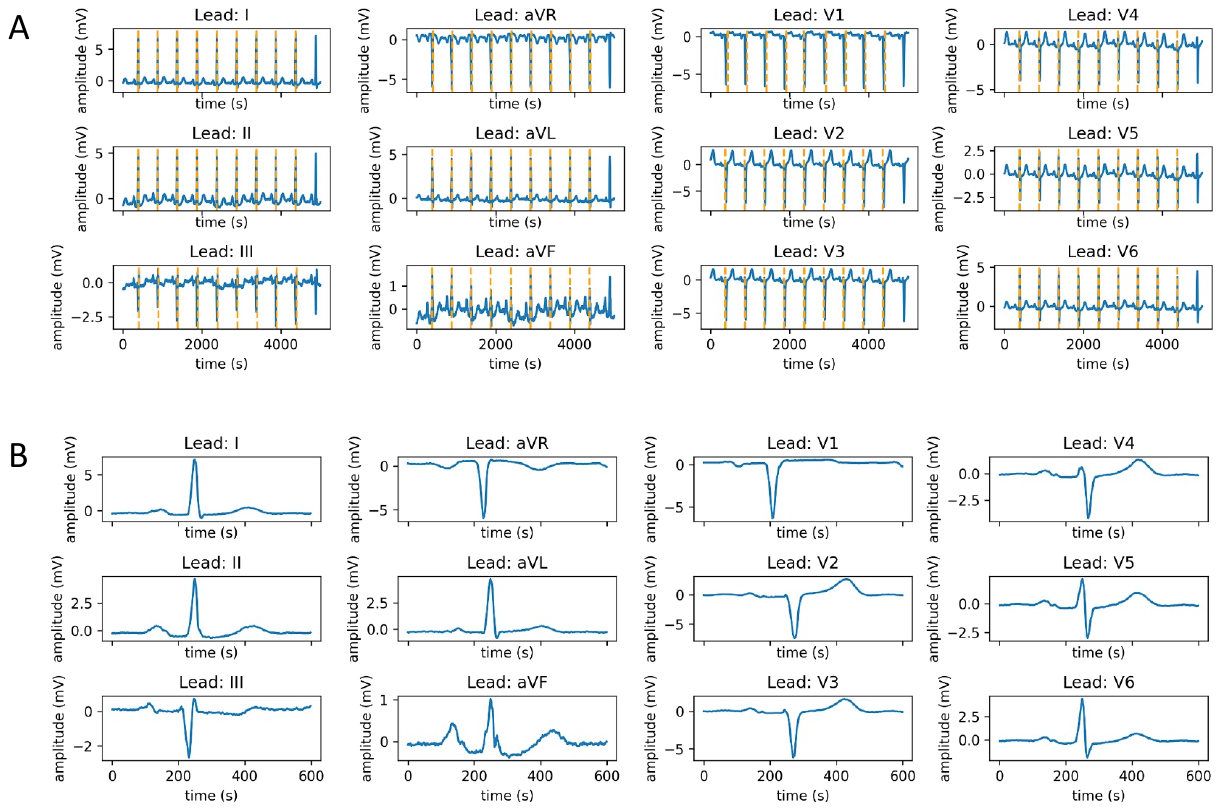

### **Supplementary Figure 3.** Cluster centroids and disease vector derivation.

Example of disease vector derivation, in this case using sex as an exemplary phenotype. The figure shows a two-dimensional t-Distributed Stochastic Neighbor Embedding (t-SNE) scatter plot and demonstrates how sex affects the distribution of electrocardiogram (ECG) encodings in latent space of 1600 UKB participants. ECG encodings from female participants (small pink dots) and male participants (small brown dots) form distinct phenotypic clusters. The larger red and blue dots mark the centroids of these clusters, respectively. The orange arrow represents the phenotype vector for sex.

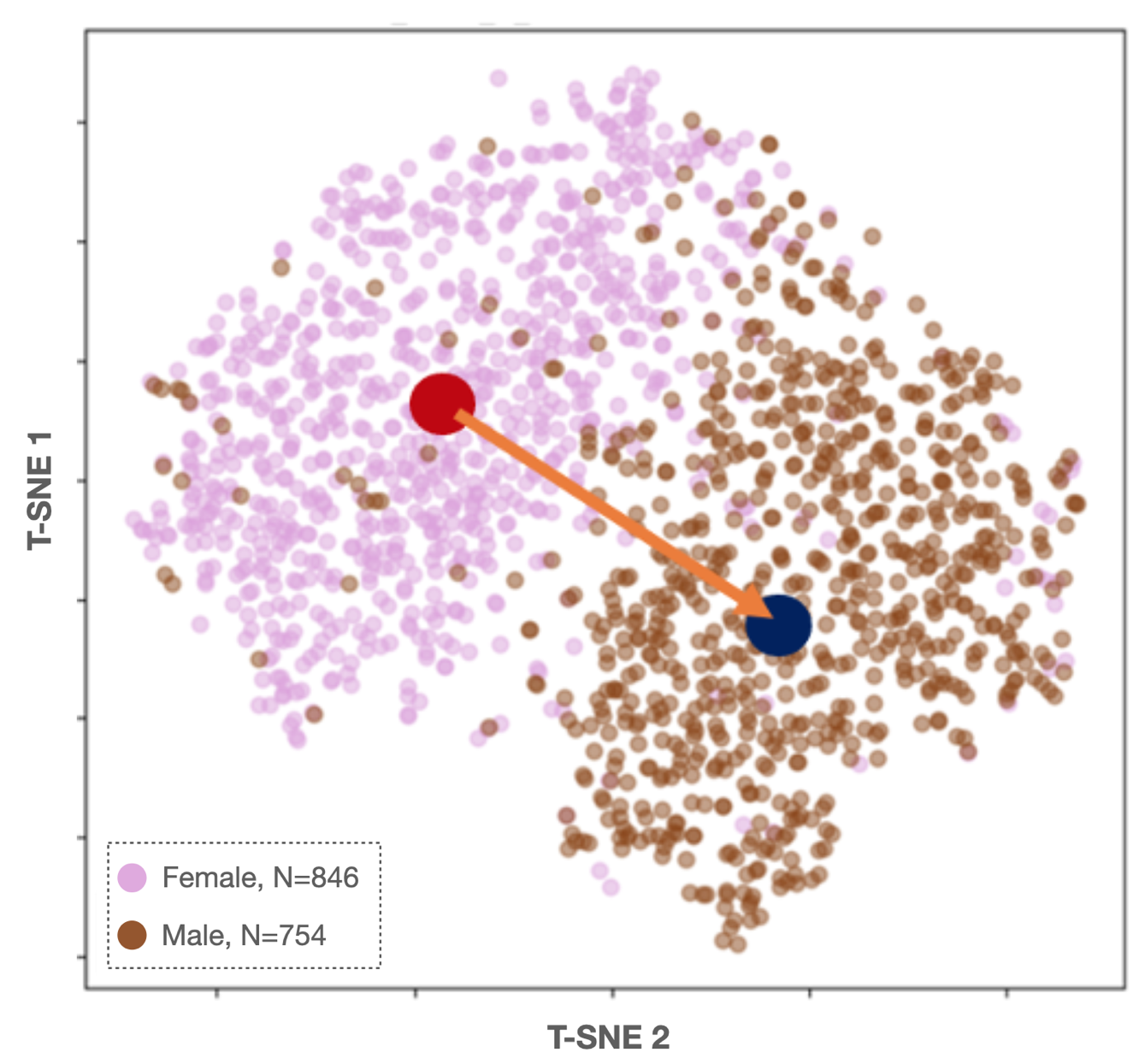

### **Supplementary Figure 4.** Latent space 12-lead electrocardiogram phenome-wide association study perturbation tests.

Panel A represents the results of the phenome-wide association study results in the Massachusetts General Hospital Community Care Cohort Project sample at baseline. Each disease tested for association is represented as a single point on the plot. The x-axis represents the phenotype category and the y-axis represents the -log10(p-value) for the association test. The remaining panels represent the results in which the disease labels are randomly reclassified with (B) 10%, (C) 20%, and D (100%) reclassification.

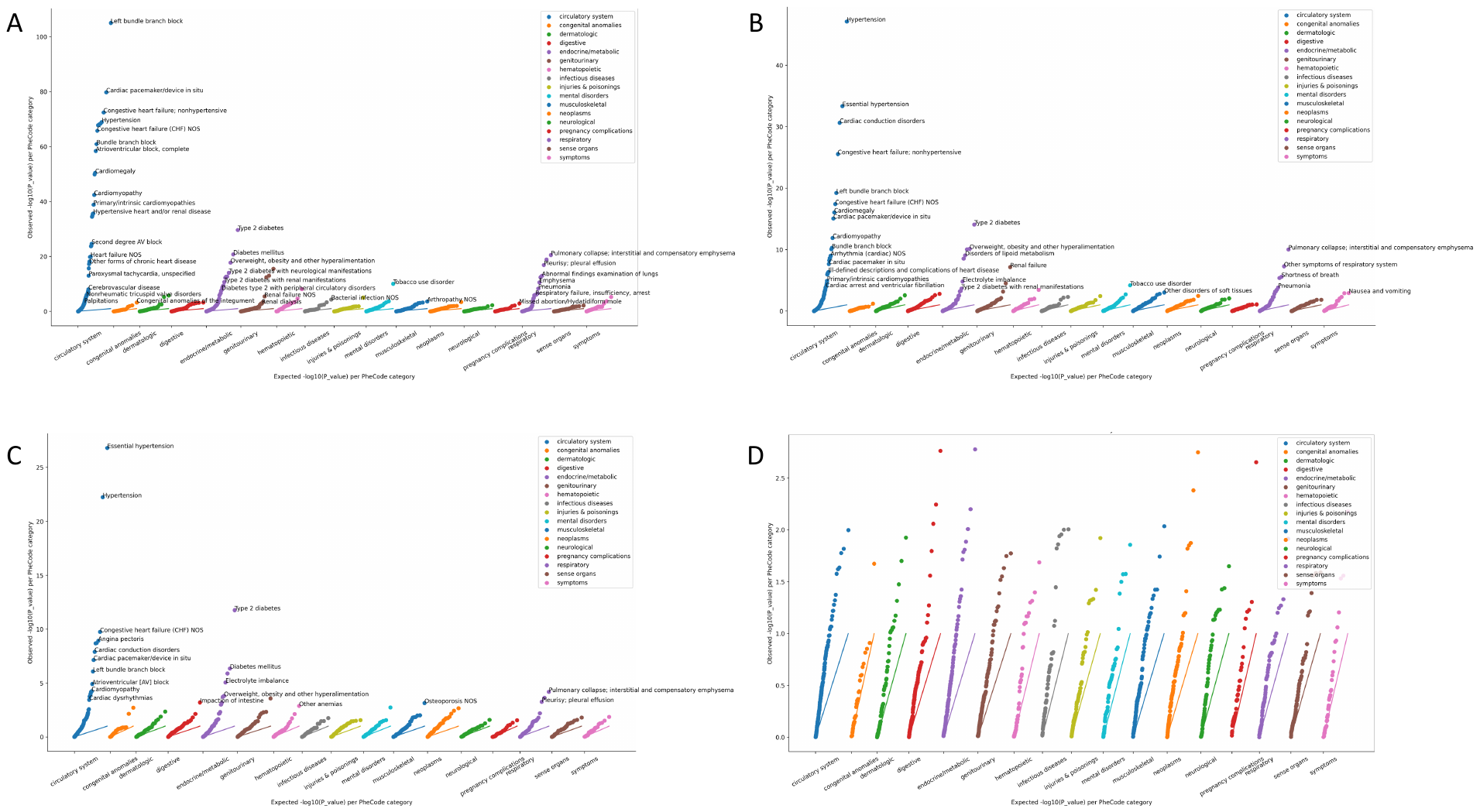

### **Supplementary Figure 5.** Forest plots demonstrating associations for the top 3 diseases in each of the datasets for both 12-lead and single-lead ECG latent space PheWAS analyses.

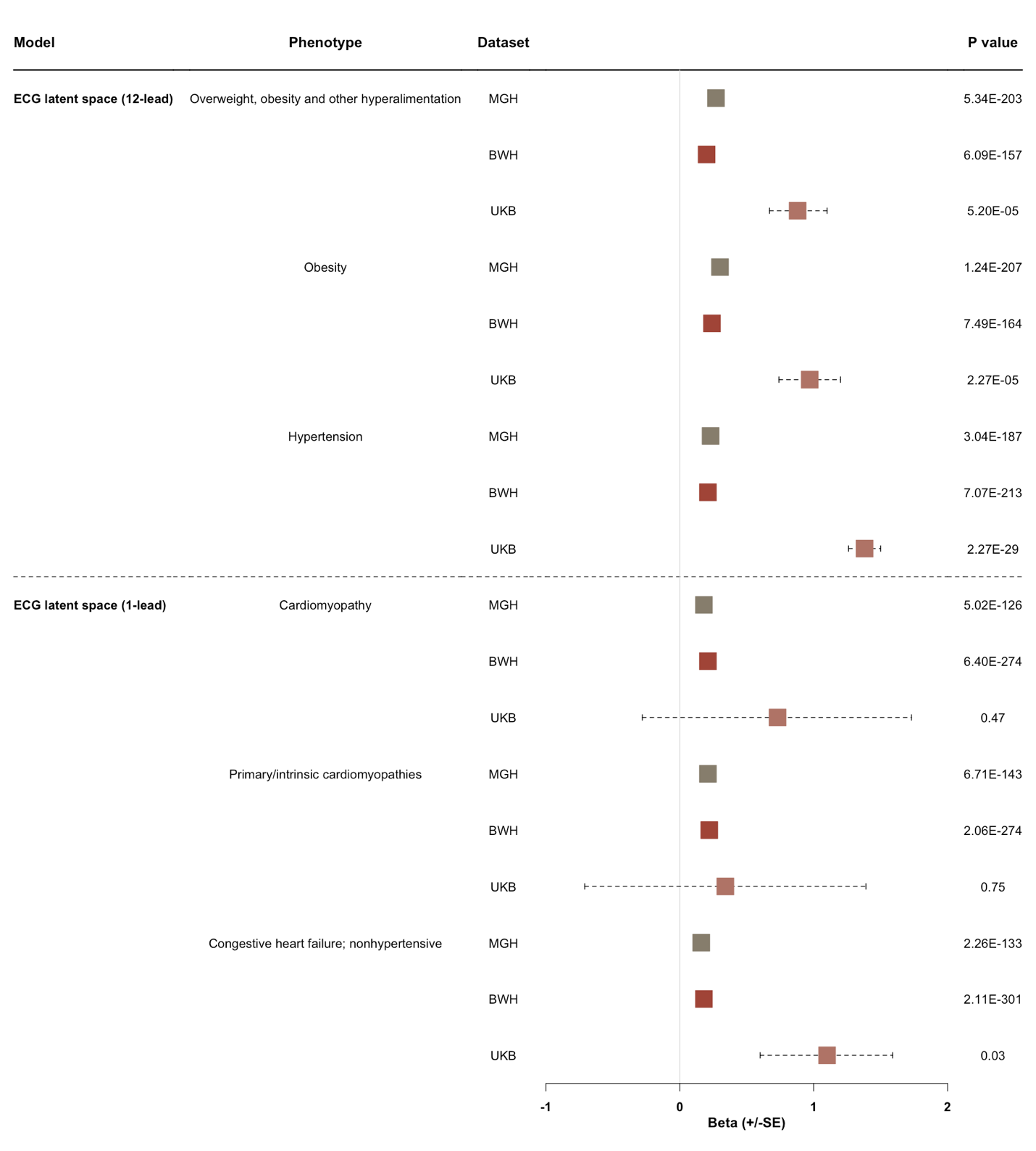

### **Supplementary Figure 6.** Discrimination of diseases using ECG projected components.

Distributions of the area under the receiver operating characteristic curves (AUC) for models with (panels A-C) and without (panels D-F) the ECG projected component for each significantly associated (p<3.1x10^-5^) disease are displayed, stratified by disease category and dataset (rows). Distributions of the delta AUC between models for each disease are displayed in panels G-I. Boxplots display the median and interquartile range, with whiskers extending to 1.5 times the interquartile range. AUCs were derived from logistic regression models adjusted for age, sex, race, and ECG projected component. In MGH-C3PO and BWH-C3PO, models were also adjusted for ECG acquisition date and zero padding (see methods).

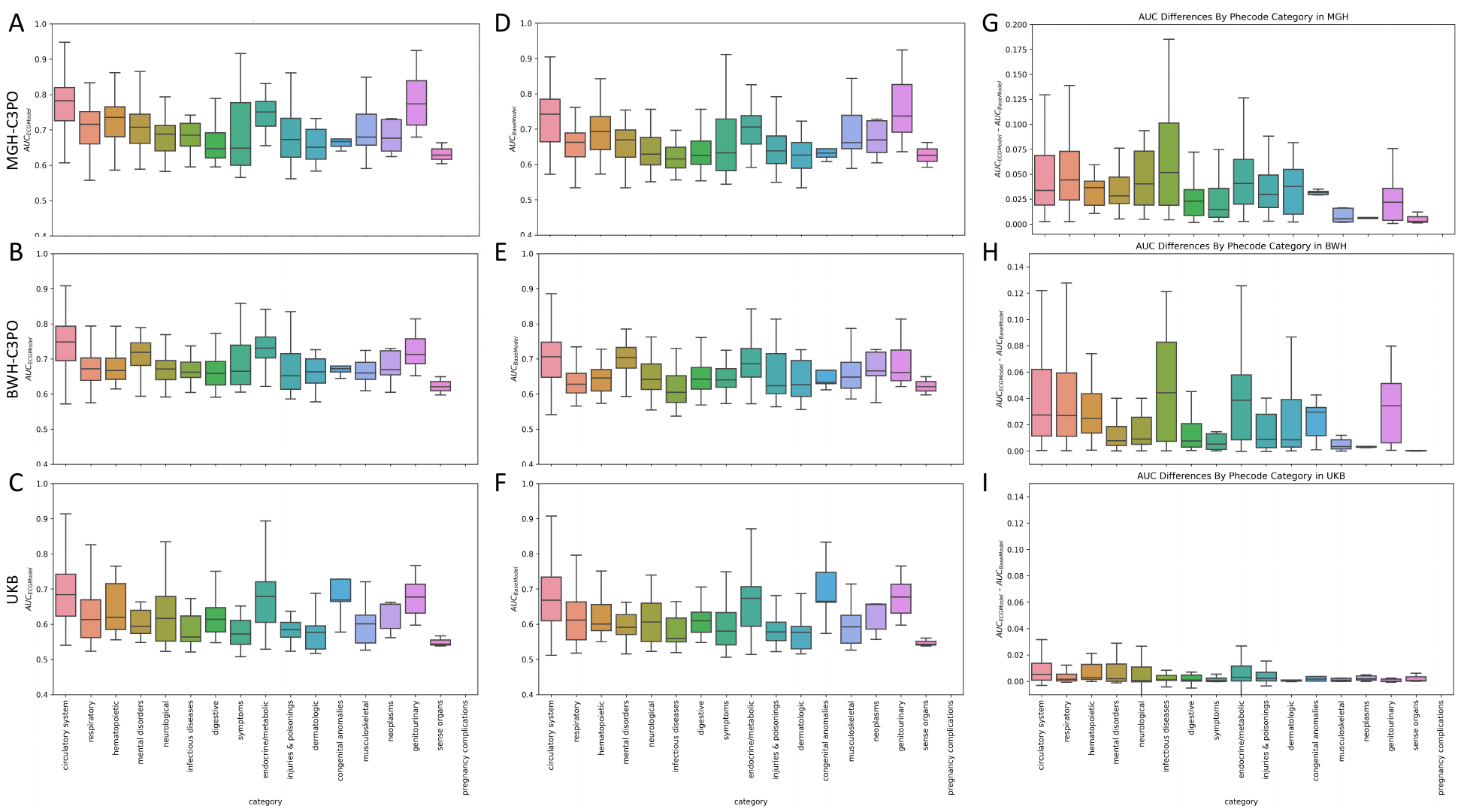

### **Supplementary Figure 7.** Disease vector correlation matrix.

Correlation matrix for disease vectors based on the 12-lead electrocardiogram model, organized by disease category. The color scale represents Pearson correlation coefficients.

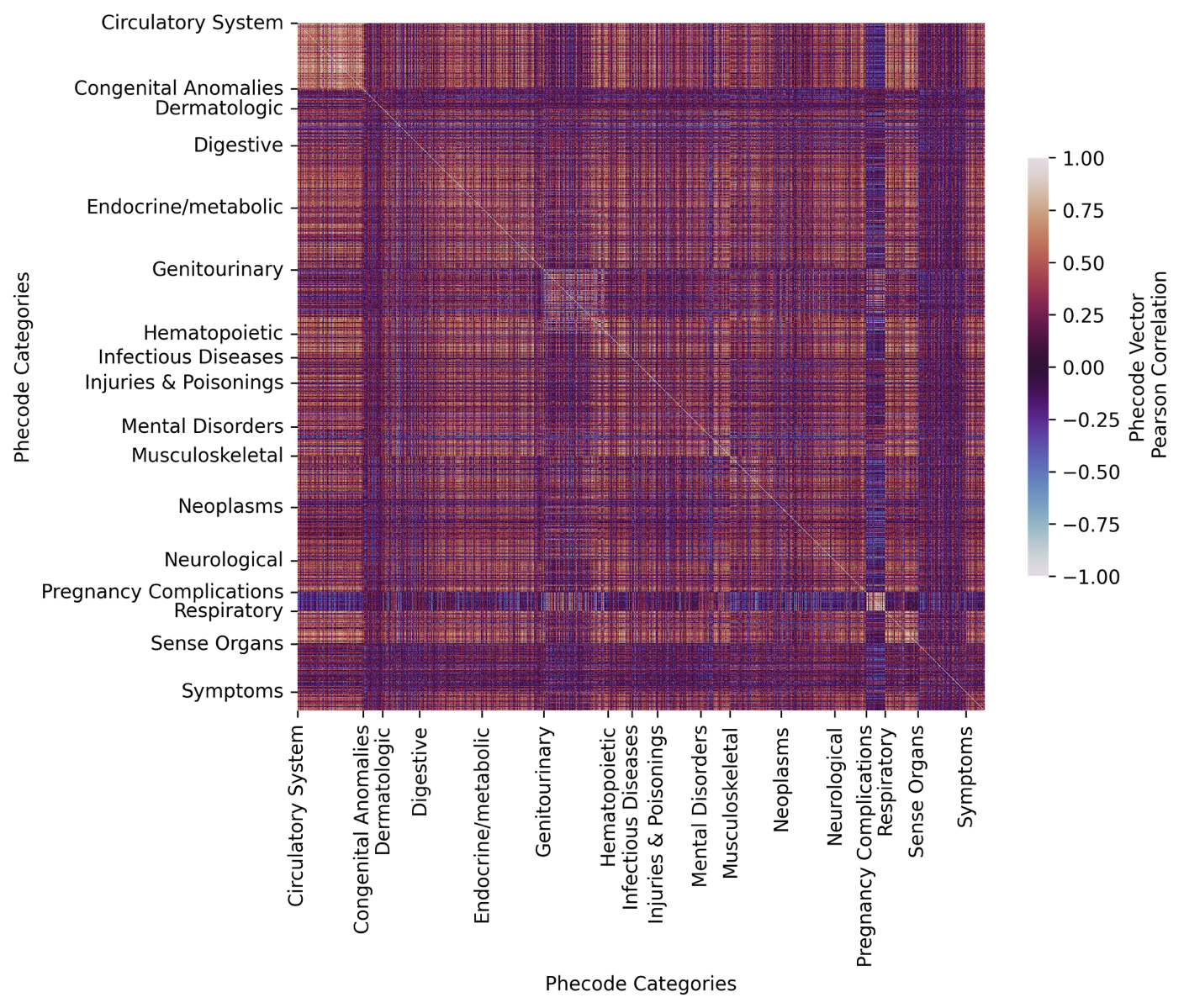

### **Supplementary Tables 1–6.**

See separate data file.
